# A retrospective comparison of deep learning to manual annotations for optic disc and optic cup segmentation in fundus photos

**DOI:** 10.1101/2020.05.05.20091660

**Authors:** Huazhu Fu, Fei Li, Yanwu Xu, Jingan Liao, Jian Xiong, Jianbing Shen, Jiang Liu, Xiulan Zhang, for iChallenge-GON study group

## Abstract

**Purpose:** Optic disc (OD) and cup (OC) segmentation are fundamental for fundus image analysis. Manual annotation is time consuming, expensive, and highly subjective, while an automated system is invaluable to the medical community. The aim of this study is to develop a deep learning system to segment OD and OC in fundus photos, and evaluate how the algorithm compares against manual annotations.

**Methods:** A total of 1200 fundus photos with 120 glaucoma cases were collected. The OD and OC annotations were labeled by seven licensed ophthalmologists, and glaucoma diagnoses were based on comprehensive evaluations of the subject medical records. A deep learning system for OD and OC segmentation was developed. The performances of segmentation and glaucoma discriminating based on the cup-to-disc ratio (CDR) of automated model were compared against the manual annotations.

**Results:** The algorithm achieved an OD dice of 0.938 (95% confidence interval (CI), 0.934-0.941), OC dice of 0.801 (95% CI, 0.793-0.809), and CDR mean absolute error (MAE) of 0.077 (95% CI, 0.073-0.082). For glaucoma discriminating based on CDR calculations, the algorithm obtained an area under receiver operator characteristic curve (AUC) of 0.948 (95% CI, 0.920-0.973), with a sensitivity of 0.850 (95% CI, 0.794-0.923) and specificity of 0.853 (95% CI, 0.798-0.918).

**Conclusions:** We demonstrated the potential of the deep learning system to assist ophthalmologists in analyzing OD and OC segmentation and discriminating glaucoma from non-glaucoma subjects based on CDR calculations.

**Translational Relevance:** We investigate the segmentation of OD and OC by deep learning system compared against the manual annotations.

## INTRODUCTION

Glaucoma is the leading cause of irreversible blindness around the world^1^. In clinical practice, glaucoma is diagnosed by evaluating the thickness of the retinal nerve fiber layer (RNFL), and the morphology of the optic nerve head (ONH)^2,3^. Some other features are considered when making a diagnosis of glaucoma^1,4^, including visual field, intraocular pressure, family history, corneal thickness, history of disc hemorrhages, etc. In fundus examinations, glaucoma is usually characterized by a larger cup-to-disc ratio (CDR), focal notching of the neuroretinal rim, etc.^5,6^. An enlarged CDR may also indicate the existence of other ocular ailments, such as neuro-ophthalmic diseases. Previous studies have shown that a larger vertical CDR is closely associated with the progression of glaucoma^7–9^. However, calculations for the CDR often vary among ophthalmologists and are relatively subjective, since they require a comprehensive judgment of shapes and structures of optic disc (OD) and optic cup (OC)^10,11^. As such, several tools incorporating computer vision and machine learning techniques have been developed to perform automated OD and OC segmentation for large-scale data analysis.

Recently, deep learning techniques have been shown to perform very well in a wide variety of medical imaging tasks^12,13^, including diabetic retinopathy screening^14–16^, and age-related macular degeneration detection^17–19^. Automated glaucoma detection from fundus photos has also received increasing attention^20–23^. However, most of the studies have focused on predicting glaucoma directly from the fundus photos, without any visualization result. By contrast, OD and OC segmentation could be helpful for calculating the risk factors (e.g., CDR, Rim-to-Disc Ratio^24^), and providing a segmentation visualization result. Moreover, although some automated segmentation methods appear to perform well on the small datasets^25–28^, they have not been compared to performance by the practicing ophthalmologists.

In this study, we developed a deep learning system for automated OD and OC segmentation in fundus photos and evaluated its performances compared against seven ophthalmologists for OD and OC segmentation and glaucoma discriminating based on CDR calculations.

## METHODS

### Data acquisition

The fundus photos were collected from Zhongshan Ophthalmic Center, Sun Yat-sen University, China. The fundus photos were captured by using Zeiss Visucam 500 and Canon CR-2 machines. We included 1496 fundus photos from 748 subjects. As long as the diagnosis of both eyes are determined, both eyes of the same subjects were included. After a quality assessment, the low-quality fundus photos (e.g., low-contrast, blurry) are excluded. Finally, a total of 1200 fundus photos are selected in our study with 120 glaucoma and 1080 non-glaucoma cases (Inclusion criteria: 1. Age ≥ 18 years old; 2. Clear images without artifacts or overexposure; 3. Definite diagnoses acquired). Diagnoses were based on the comprehensive evaluation of the subjects’ medical records, including fundus photos, IOP measurements, optical coherence tomography images, visual fields (VF). The fundus photos came from previous clinical studies^29^, and all the participants signed informed consent before enrollment. IRB/Ethics Committee ruled that approval was not required for this study.

The dataset was split into a training set (400 photos with 40 glaucoma cases), a validation set (400 photos with 40 glaucoma cases, female: 52 %, mean age: 25.3 ± 11.5), and a test set (400 photos with 40 glaucoma cases, female: 55 %, mean age: 23.7 ± 9.0), following the REFUGE challenge^29^. The photos from the same patient were assigned to the same set. The training set was used to learn the algorithm parameters, the validation set was used to choose model, and the test set was used to evaluate the algorithm, as well as the ophthalmologists.

### Diagnostic criteria for glaucoma

Patients with glaucomatous damage in the ONH area and reproducible glaucomatous VF defects were included in our study. A glaucomatous VF defect is defined as a reproducible reduction in sensitivity compared to the normative dataset, in reliable tests, at: (1) two or more contiguous locations with p-value < 0.01, (2) three or more contiguous locations with p-value < 0.05. ONH damage is defined as CDR > 0.7, thinning of RNFL (an RNFL defect in the optic nerve head shown on the OCT reports), or both, without a retinal or neurological cause for VF loss. Specifically, First, the diagnostic criteria was based on the trial in glaucoma, i.e., UKGTS^30^. Second, if the points exist on the rim, there could be false-positive cases. However, as mentioned in our manuscript, the included subjects in our study received repeated VF tests to ensure reliability. If the defects exist all the time, we consider them as glaucomatous defects.

All OD and OC annotations were manually labeled by seven licensed ophthalmologists (average experience: 8 years, range: 5-10 years). All ophthalmologists independently reviewed and marked OD and OC in each photo as the tilted ellipses using a free image labelling tool with capabilities for image review, zoom, and ellipse fitting. Ophthalmologists did not have access to any patient information or knowledge of disease prevalence in the data. The final standard reference labels of OD and OC were created by merging the annotations from multiple ophthalmologists using majority voting. Specifically, a senior specialist with more than 10 years of experience in glaucoma performed a quality check afterward, analyzing the resulting masks to account for potential mistakes. When errors in the annotations were observed, this additional reader analyzed each of the seven segmentations, removed those that were considered failed in his/her opinion and repeated the majority voting process with the remaining ones. Only a few cases had to be corrected using this protocol.

### Algorithm development

In this study, we proposed a deep learning system for automated OD and OC segmentation in fundus photos (Figure 1). The proposed system included two main stages: (1) OD region detection, which first localized the OD center within the whole fundus photo, and then cropped the OD region to remove the background; and (2) OD and OC segmentation, which segmented the OD and OC jointly via a multi-label deep network in the cropped OD image. We employed a U-Net network for OD region detection, which was based on encoder–decoder architecture to achieve satisfactory performances in many biomedical image tasks^31^. The encoder path consisted of the multiple convolutional layers with various filter banks to produce a set of feature representations for the inputs, while the decoder path aggregated the feature representations to predict the probability map of the OD region in the fundus photo. Additionally, skip connections were used to concatenate the feature representations from the encoder path to the corresponding decoder path. The final output of the U-Net network was a probability map, indicating the OD region and background for each pixel in the fundus image, as shown as Figure 1 (c). The implementation details of the U-Net network for OD detection were given in the Supplement A. With the probability map of OD localization, we used a thresholding of 0.5 to obtain the mask for the OD region, and cropped a local image around the OD for the following OD and OC segmentation stage.

**Figure 1.**
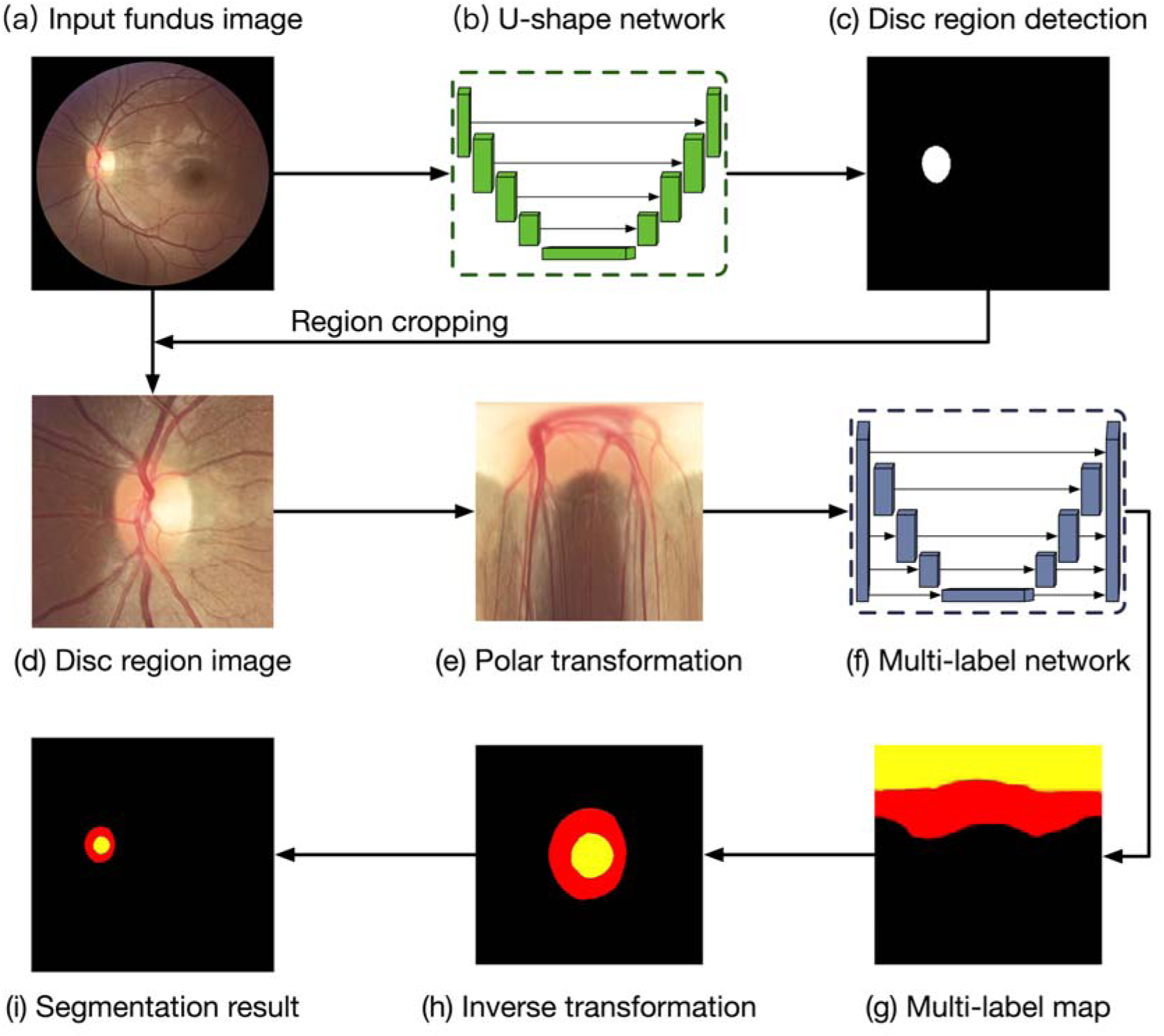
Deep learning system for automated segmentation in fundus images. The algorithm included two stages: Optic disc (OD) region detection and OD and optic cup (OC) segmentation. For a given fundus image (a), the U-Net network (b) was utilized to detect the OD region (c). With the cropped OD region (d), a polar transformation was used to map the image into polar coordinate (e). A multi-label network (f) segmented the OD and OC jointly, and an inverse transformation returned the output map (g) back to original coordinates (h).

In the second stage of our algorithm, a multi-label network was utilized to segment OD and OC simultaneously in the cropped OD region image^26^. Similar to the U-Net network, the multi-label network also consisted of an encoder and a decoder path based on convolutional layers. The difference is that the multi-label network employed the average pooling layers to naturally down-sample the images as multi-scale inputs to the corresponding encoder path, while the multi-scale outputs from each scale of decoder path were fused together as the final probability map. Additionally, the multi-label loss function was used to learn the binary classifier of each class (i.e., OD and OC), and assign multiple labels to each pixel for segmentation of OD and OC jointly. The implementation details of the multi-label network for OD and OC segmentation were given in the Supplement B. In the fundus photo, the size ratio of OC region is less than the OD and background, which could lead overfitting of deep model during training. To address this, we map the OD region image into the polar coordinates, before being fed into the multi-label network. Polar transformations were carried out using the OD center as the origin and the local image width as the radius (see Figure 1 (e)). The implementation details of polar transformations were given in the Supplement C. After passing through the multi-label network, an inverse polar transformation reverted the predicted map back to the original coordinates.

The U-Net network for OD detection and multi-label network for OD and OC segmentation were trained separately. The U-Net network was trained based on the whole fundus images resized to 800 by 800 pixels, with the OD reference label, while the multi-label network was trained based on the OD region images resized to 400 by 400 pixels, with the OD and OC reference labels. Random flips and rotations were applied to all training photos before they were fed into the networks for data augmentation. These two networks were implemented with Python (version 3.6) based on Keras (version 2.2) with a Tensorflow (version 1.12) backend. All network parameters of the networks were optimized by using stochastic gradient descent with a learning rate of 0.0001 and a momentum of 0.9. In order to prevent the networks from overfitting, early stopping was performed, which saved the network model after each epoch and chose the final model with the lowest loss on the validation set. Each stage of training required around 2 hours for completion, on a single NVIDIA Titan XP.

### Statistical analysis and evaluation

For segmentation evaluation, we reported three performance metrics, namely, OD dice, OC dice, and CDR mean absolute error (MAE). The dice scores measured the overlap ratio between the target regions of the reference label and segmented result, while CDR MAE was the mean absolute error between the calculated CDR values from the reference label and segmented result. We also determined the standard deviation (SD) and 95% Bayesian confidence interval (CI)^32^ for each segmentation metric.

In addition to evaluating the segmentation performance, we also compared the algorithm against ophthalmologists for discriminating glaucoma from non-glaucoma photos based on CDR calculations. The performances across different diagnostic thresholds of CDR were assessed in terms of the area under receiver operator characteristic curve (AUC). To convert the CDR to a binary prediction, we chose the highest point on the ROC curve, which offers minimal trade-off between sensitivity and specificity, as the final discriminating threshold. Moreover, the 95% bootstrapping CI^33^ was provided for each discriminating metric as: computing 10,000 bootstrap replicates from the set, and each metric was computed for algorithm and reference label on the same bootstrap replicate. p-values were reported by comparing the AUC with the algorithm and ophthalmologist predictions. All statistical analyses were performed using Python (version 3.6) with SciPy (version 1.2) and Scikit-learn (version 2.20). Figures were created using Matplotlib (version 3.0) and Seaborn (version 0.9).

## RESULTS

The segmentation performances of our algorithm and annotations of the seven ophthalmologists, for the test set, were listed in Table 1. For glaucoma data, the algorithm obtained an OD dice of 0.941 (SD, 0.057, 95% CI, 0.926-0.956), OC dice of 0.864 (SD, 0.089, 95% CI, 0.841-0.887), and CDR MAE of 0.065 (SD, 0.056, 95% CI, 0.051-0.080). For non-glaucoma data, the algorithm predicted an OD dice of 0.937 (SD, 0.040, 95% CI, 0.934-0.941), OC dice of 0.794 (SD, 0.096, 95% CI, 0.786-0.803), and CDR MAE of 0.079 (SD, 0.050, 95% CI, 0.074-0.083). The segmentation performances of the algorithm on the whole test set achieved an OD dice of 0.938 (SD, 0.041, 95% CI, 0.934-0.941), OC dice of 0.801 (SD, 0.097, 95% CI, 0.793-0.809), and CDR MAE of 0.077 (SD, 0.051, 95% CI, 0.073-0.082). For OD segmentation, the algorithm performed better than Ophthalmologist 2, who reported an OD dice of 0.928 (SD, 0.046, 95% CI, 0.925–0.932), and Ophthalmologist 3, who determined the OD dice to be 0.924 (SD, 0.039, 95% CI, 0.921-0.927). For OC segmentation, the algorithm performed better than Ophthalmologist 1, who obtained an OC dice of 0.705 (SD, 0.121, 95% CI, 0.695-0.715), and Ophthalmologist 7, who got an OC dice of 0.670 (SD, 0.138, 95% CI, 0.658-0.681). The OD and OC dice scores of inter-agreement for seven ophthalmologists were given in Figure 2. Boxplots for the calculated CDRs of the reference label, the ophthalmologist annotations and the algorithm outputs, for glaucoma and non-glaucoma data on test set, were plotted in Figure 3. The average CDRs of the reference labels for the glaucoma and normal cases were 0.656 and 0.453, respectively for test set.

**Table 1:**
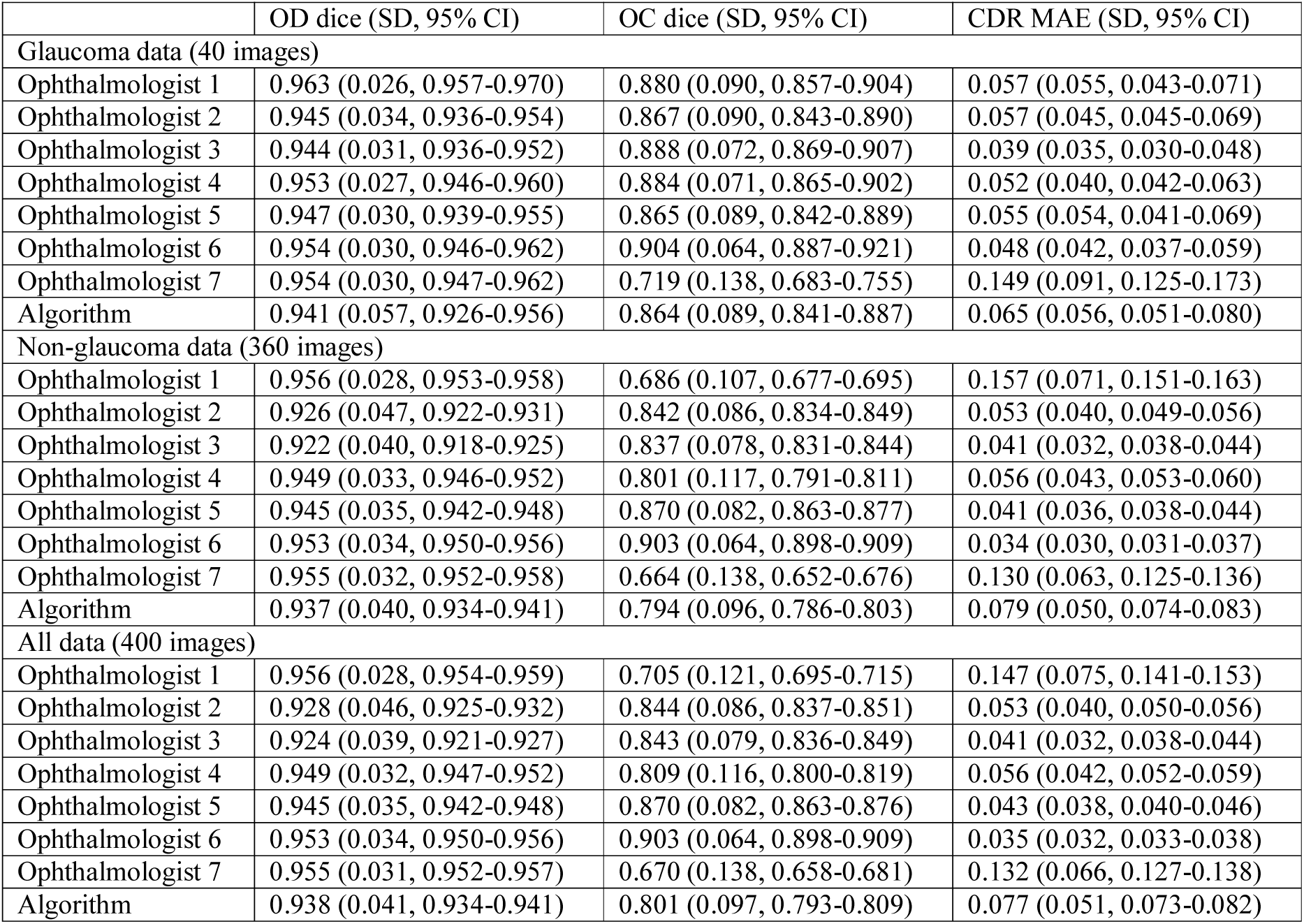
Segmentation performances of ophthalmologists and algorithm on test set. **OD** = optic disc; **OC** = optic cup; **CDR** = cup-to-disc ratio; **MAE** = mean absolute error; **CI** = confidence interval; **SD** = standard deviation.

**Figure 2.**
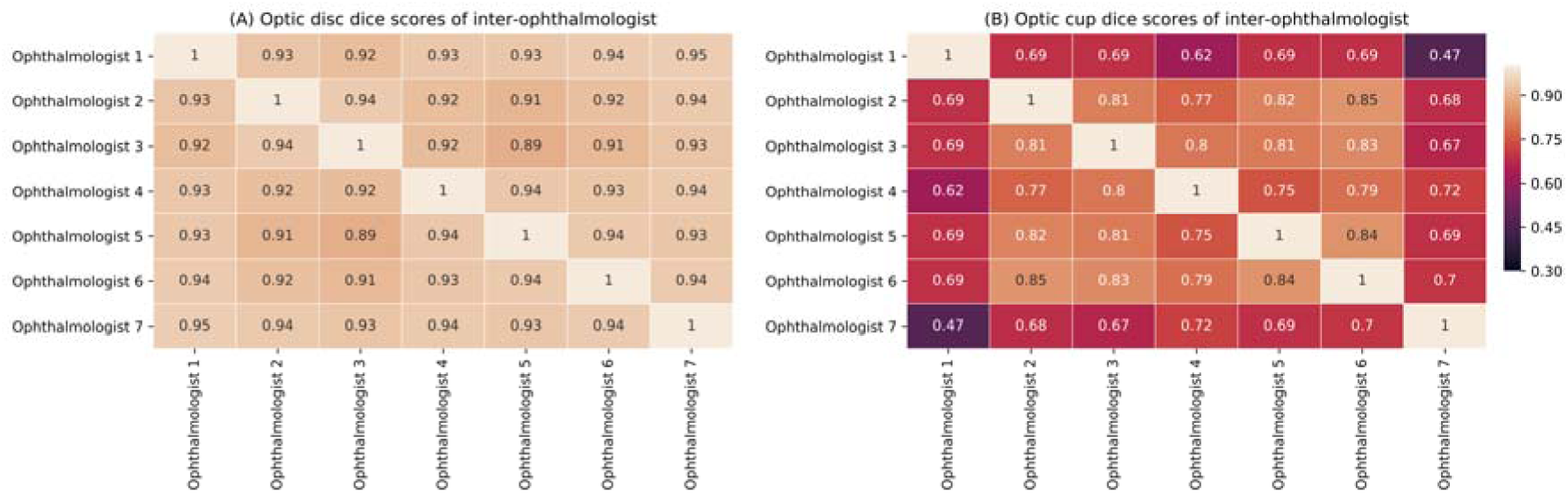
Dice scores of inter-agreement for seven ophthalmologists on the test set for (A) optic disc, and (B) optic cup.

**Figure 3.**
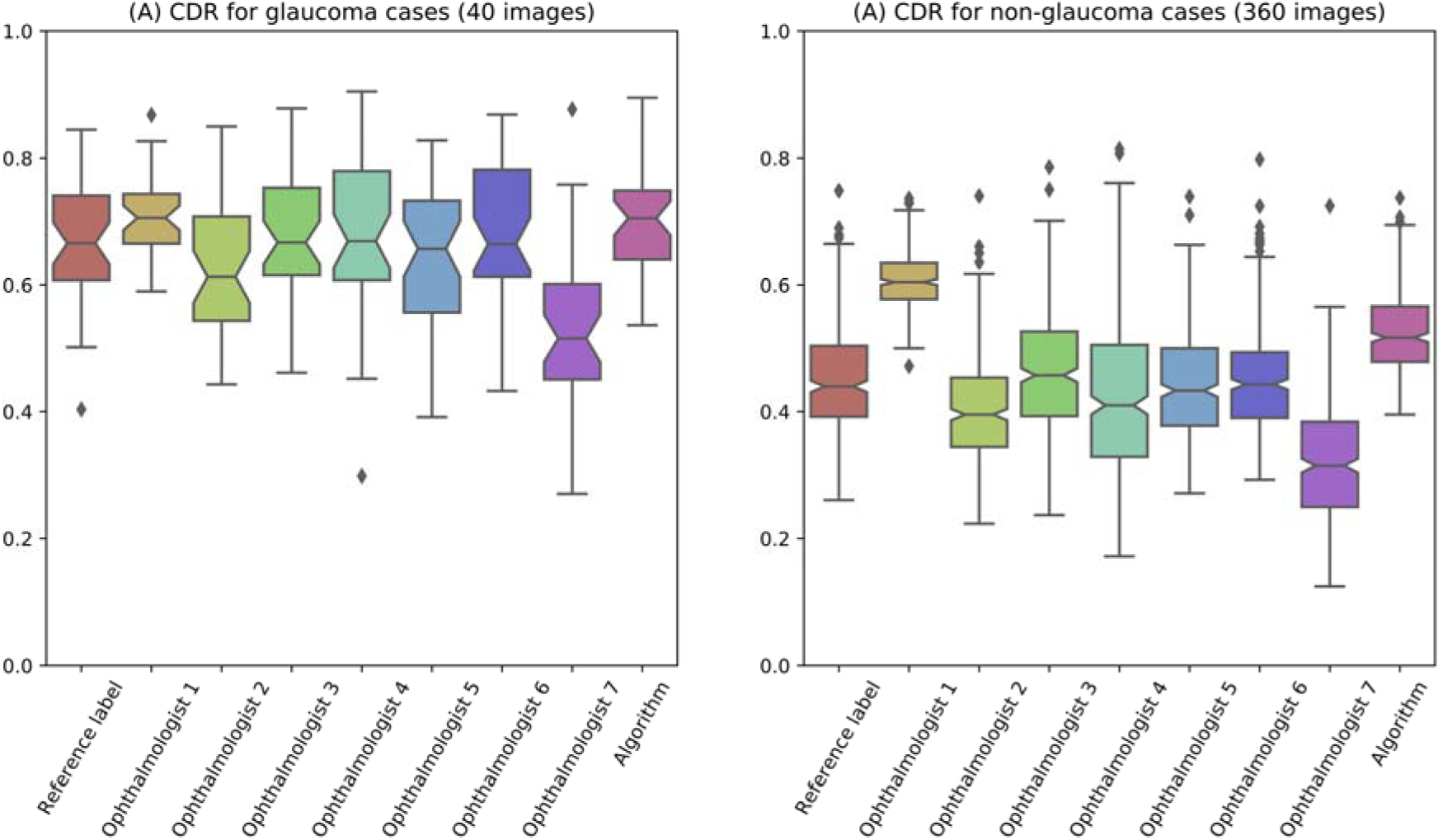
Boxplots of the calculated cup-to-disc ratio (CDR) from segmentation results on test set.

Figure 4 showed the visual results of automated OD and OC segmentation, for both glaucoma and non-glaucoma data. Several failure cases were also provided in Figure 4 (E, F). One common failure case for OD segmentation was confusion when peripapillary atrophy (PPA) was present, since this looks similar to the OD (green arrow in Figure 4 (E)). Failure cases also occurred due to the low-quality of the fundus photos, where poor illumination and low-contrast often made it difficult to determine the boundary of the OC (green arrow in Figure 4 (F)). However, this could be relieved using additional image enhancement pre-processing.

**Figure 4.**
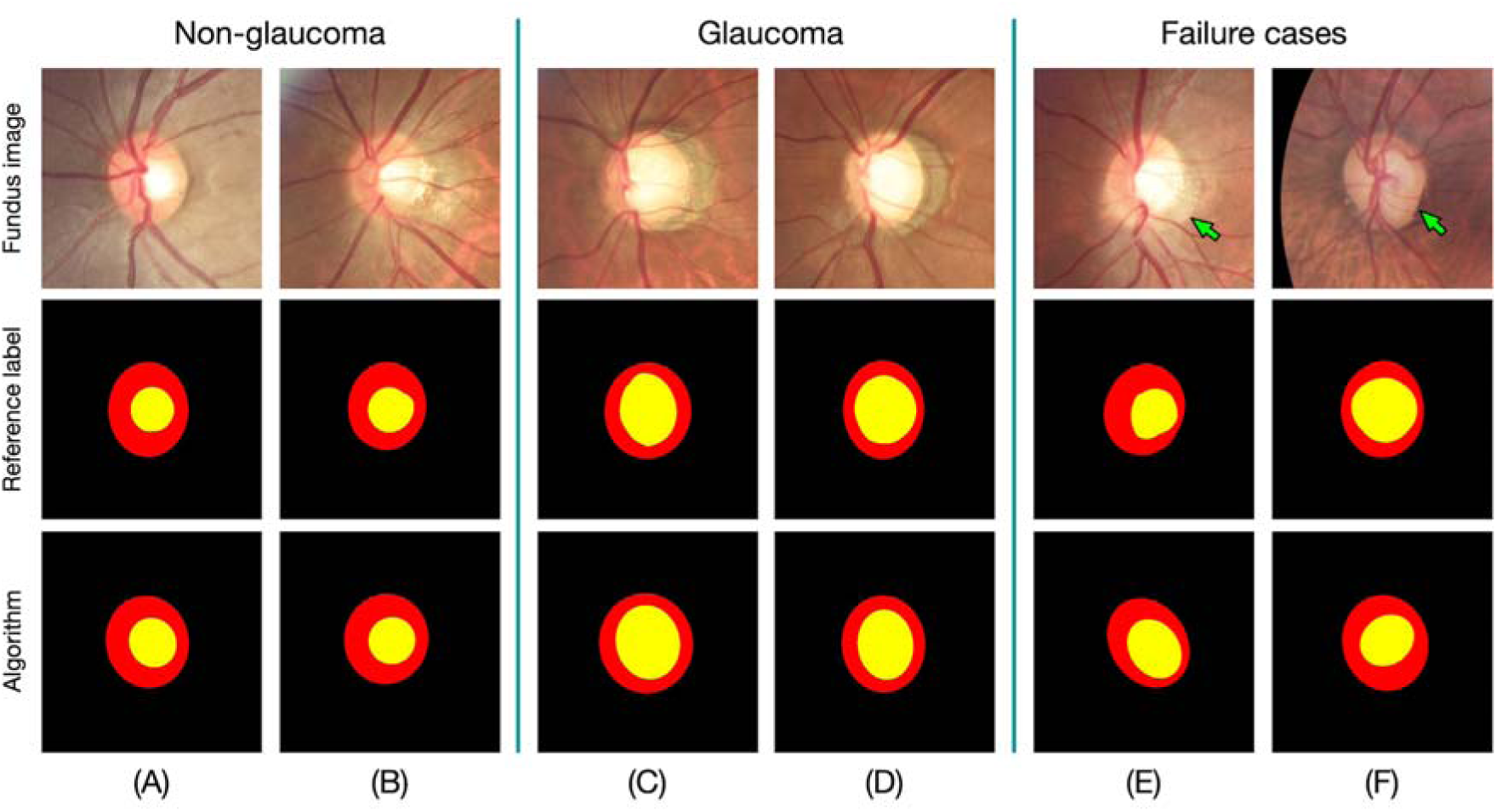
The visual results of segmentation. The segmented optic disc and optic cup regions were labeled by red and yellow colors, respectively. (a, b) Glaucoma cases, (c, d) non-glaucoma cases, (e, f) failure cases.

The performances of discriminating glaucoma from non-glaucoma subjects based on CDR, for test set, were shown in Figure 5 and Table 2. The algorithm obtained an AUC of 0.948 (95% CI, 0.920-0.973), with a sensitivity of 0.850 (95% CI, 0.794–0.923) and specificity of 0.853 (95% CI, 0.798-0.918). The algorithm obtained the rank 2 discriminating performance, only lower than ophthalmologist 2, who got an AUC of 0.956 (95% CI, 0.933-0.975, p-value < 0.0001). Moreover, Figure 4 (E) and (F) showed the false negative and false positive samples, respectively.

**Figure 5.**
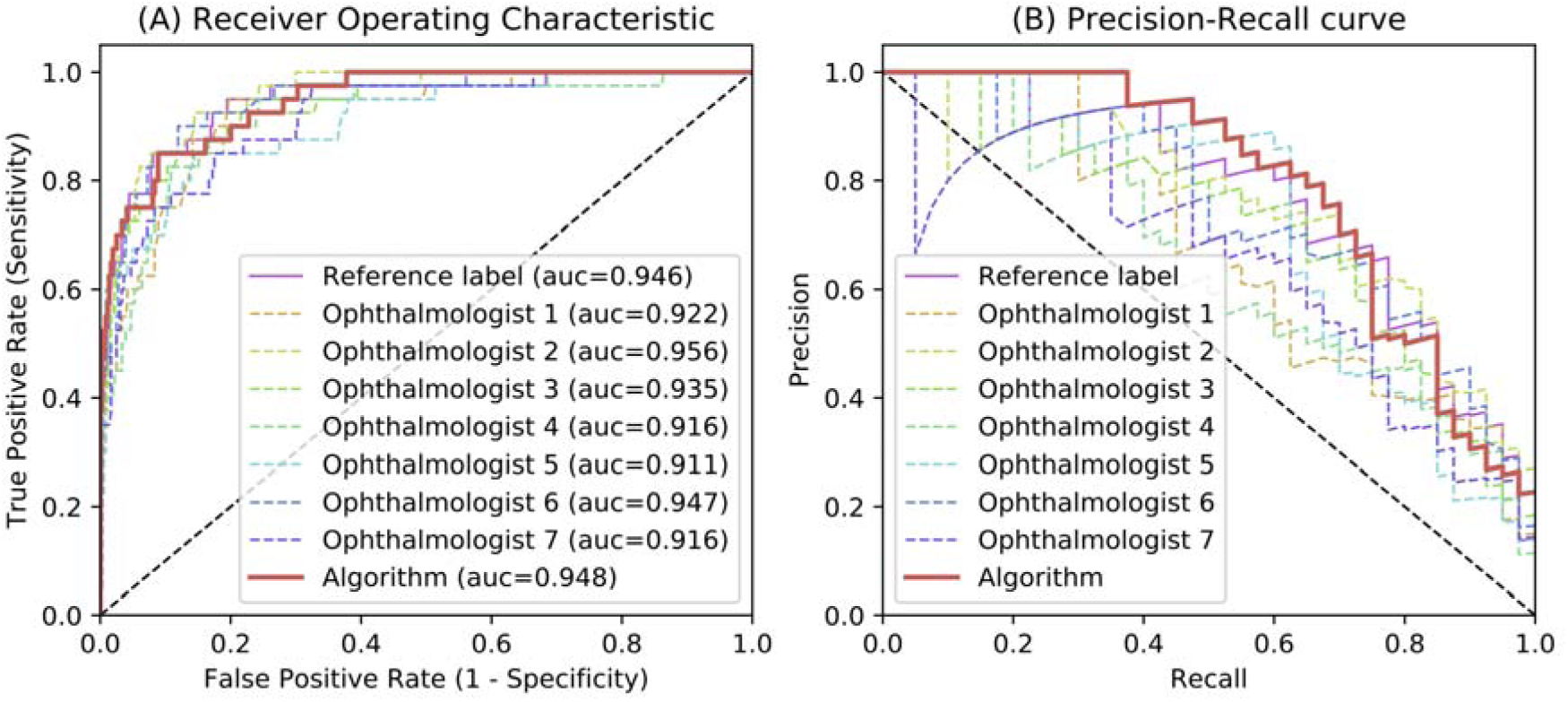
(A) The average receiver operating characteristic curves (AUC) for glaucoma diagnosis based on cup to disc ratio (CDR) on test set. (B) The precision-recall curves for glaucoma diagnosis based on cup to disc ratio (CDR) on test set.

**Table 2:** Diagnosis performances of experts and algorithm on test set. **AUC** = area under the eceiver operating characteristic curve; **CI** = confidence interval.

|  | AUC (95% CI) | p-value | Sensitivity (95% CI) | Specificity (95% CI) | Precision (95% CI) |
| --- | --- | --- | --- | --- | --- |
| Reference label | 0.946 (0.911-0.974) | < 0.0001 | 0.875 (0.811-0.927) | 0.867 (0.819-0.923) | 0.700 (0.588-0.791) |
| Ophthalmologist 1 | 0.922 (0.884-0.955) | < 0.0001 | 0.875 (0.800-0.897) | 0.858 (0.812-0.887) | 0.585 (0.464-0.694) |
| Ophthalmologist 2 | 0.956 (0.933-0.975) | < 0.0001 | 0.875 (0.826-0.930) | 0.869 (0.834-0.928) | 0.700 (0.577-0.786) |
| Ophthalmologist 3 | 0.935 (0.902-0.964) | < 0.0001 | 0.850 (0.789-0.913) | 0.850 (0.795-0.908) | 0.650 (0.556-0.771) |
| Ophthalmologist 4 | 0.916 (0.871-0.954) | < 0.0001 | 0.850 (0.791-0.903) | 0.847 (0.796-0.898) | 0.575 (0.438-0.667) |
| Ophthalmologist 5 | 0.911 (0.866-0.951) | < 0.0001 | 0.850 (0.756-0.892) | 0.856 (0.755-0.886) | 0.625 (0.525-0.755) |
| Ophthalmologist 6 | 0.947 (0.916-0.972) | < 0.0001 | 0.900 (0.824-0.930) | 0.881 (0.832-0.922) | 0.659 (0.556-0.773) |
| Ophthalmologist 7 | 0.916 (0.876-0.951) | < 0.0001 | 0.825 (0.765-0.882) | 0.825 (0.772-0.878) | 0.625 (0.469-0.720) |
| Algorithm | 0.948 (0.920-0.973) | - | 0.850 (0.794-0.923) | 0.853 (0.798-0.918) | 0.700 (0.600-0.800) |

## DISCUSSION

The purpose of this study was to develop a deep learning algorithm for automated OD and OC segmentation in fundus photos and compare its performance to ophthalmologist annotations. The results demonstrated that the proposed deep learning algorithm achieved satisfactory performances on the OD and OC segmentation task and the glaucoma discriminating task based on CDR calculations.

OD and OC segmentation are fundamental for fundus analysis, especially for CDR calculations during discriminating glaucoma from non-glaucoma subjects. Developing an automated system for this task is crucial. First, as briefly mentioned, manual fundus photo labelling is highly time-consuming, with the average ophthalmologist requiring 40 seconds to annotate a single photo. Because our algorithm could reduce this time to 2 second, it would be highly beneficial for accelerating processing time and analyzing large-scale datasets. Second, manual annotations are highly subjective. In fact, the segmentations carried out by the ophthalmologists were easily affected by both fundus resolution and image quality. The inter-agreement rating between the various ophthalmologists, for both OD and OC dice scores on the test set, were shown in Figure 2. As can be seen, there was slight variation between the OD segmentation results, with inter-agreement scores ranging from 0.89 to 0.95. However, the OC segmentation task suffered a larger variability, with inter-agreement scores ranging from 0.47 to 0.85. The boundary of OD was clear and definite enough to determine in fundus photo, which produced a high inter-agreement score between the by ophthalmologists, as shown in Figure 2 (A). Different from the OD, the boundary of OC was more difficult to identify, which was influenced by many factors such as tilted disc, illumination, and low contrast, etc. These factors may result in the clinical uncertainty during different ophthalmologists and a variable OC segmentation. Moreover, OC segmentation by an ophthalmologist was a highly subjective task, which was related to individual bias and clinical experiences. This also led a low inter-agreement score (See Figure 2 (B)). By contrast, the automated algorithm provided a consistent result for the same photo with freezing the trained parameters and model. Moreover, due to limited GPU memory capabilities and parameter size constraints, input fundus photos had to be down-sampled for training, thus removing the requirement for high-resolution photos. Another observation is that the performances of algorithm on glaucoma cases (OD dice of 0.941, cup dice of 0.864, CDR MAE of 0.065) was better than its on non-glaucoma cases (OD dice of 0.937, cup dice of 0.794, CDR MAE of 0.079). One reason is that the advanced glaucoma cases with severe cupping usually present more clear interfaces between the OD and OC.

Over the decades, many automated deep learning algorithms have been proposed for glaucoma diagnosis in fundus photos^22,34^, optical coherence tomography (OCT)^35,36^, and anterior segment OCT (AS-OCT)^37,38^. However, while many of these produce diagnostic results from fundus photos directly, they lacked clinical interpretability and analyticity. By contrast, segmentation based algorithms generate a visible segmentation result and have more potential for clinical assistant and analysis. Some automated algorithms based on various visual features and machine learning techniques have been developed for segmenting OD and OC^28,39,40^. Cheng *et al*.^25^ classified each superpixel in fundus image with various hand-crafted features as OD and OC segmentation and reported an OD dice of 0.905 and OC dice of 0.759. Zheng *et al*.^41^ integrated the OD and OC segmentation within a graph-cut framework. However, they only utilized hand-crafted features, which were affected by the low quality of fundus photos. In our study, a multi-label deep network was employed to obtain highly discriminative representations and segment the OD and OC jointly with the multi-label loss. The results demonstrated that the proposed method enabled automated OD and OC segmentation with a comparable performance to ophthalmologists. We also evaluated the model for discriminating glaucoma from non-glaucoma subjects based on CDR calculations, which were calculated based on the segmentation results as an important glaucoma indicator. The proposed algorithm performed extremely well in comparison to ophthalmologists for glaucoma discriminating.

One limitation of this study was a specific Chinese population was evaluated and the results may not apply to other ethnic groups. Another potential limitation of our study was that the fundus photos were only taken using Zeiss Visucam 500 and Canon CR-2 cameras. This could possibly have a negative effect on the quality and performance when the algorithm was applied to images from other fundus acquisition devices. Third, in our study, we added CDR calculations as one of the clues for glaucoma diagnosis. However, some patients were shown to have a small CDR despite significant visual field loss, while others displayed a large CDR without reporting any VF loss^7^. The dataset contained 10% of glaucoma subjects and most of these glaucoma subjects were at moderate or advanced stage, the difficulty of discriminating glaucoma from non-glaucoma subjects based on CDR calculation was relatively lower. The performance of the algorithm may go down in another larger dataset. Future studies were needed to explore whether other annotations such as RNFL defects would further enhance the performance of the algorithm. Besides, early-stage glaucoma is very hard to diagnose through fundus photos. It would be interesting to add more photos from early-stage patients and train the algorithm to make diagnosis. We may try to find new clues other than CDR or RNFL defects in glaucoma discriminating based on fundus photos.

In summary, we developed and investigated a deep learning system for OD and OC segmentation in fundus images. Deep learning technique was shown to be a promising technology for helping clinicians to reliably and rapidly identify OD and OC regions. Moreover, we also evaluated discriminating glaucoma from non-glaucoma subjects based on the CDR calculations, where the proposed algorithm performed extremely well in comparison to ophthalmologists, obtaining an AUC of 0.946. As such, our technique shown high potential for assisting ophthalmologists in fundus analysis and glaucoma screening.

## Data Availability

https://refuge.grand-challenge.org

## Abbreviations and Acronyms

OD: optic disc
OC: optic cup
CDR: cup-to-disc ratio
MAE: mean absolute error
CI: confidence interval
AUC: area under the receiver operating characteristic curve
ONH: optic nerve head
RNFL: retinal nerve fiber layer
IOP: intraocular pressure
VF: visual fields
SD: standard deviation
OCT: optical coherence tomography
AS-OCT: anterior segment OCT

## REFERENCES

1. Jonas JB, Aung T, Bourne RR, Bron AM, Ritch R, Panda-Jonas S. Glaucoma. Lancet. 2017;390(10108):2183–2193. doi:10.1016/S0140-6736(17)31469-1

2. Medeiros FA, Zangwill LM, Bowd C, Vessani RM, Susanna R, Weinreb RN. Evaluation of retinal nerve fiber layer, optic nerve head, and macular thickness measurements for glaucoma detection using optical coherence tomography. Am J Ophthalmol. 2005;139(1):44–55. doi:10.1016/j.ajo.2004.08.069

3. Bussel II, Wollstein G, Schuman JS. OCT for glaucoma diagnosis, screening and detection of glaucoma progression. Br J Ophthalmol. 2014;98(Suppl 2):ii15-ii19. doi: 10.1136/bjophthalmol-2013-304326

4. Weinreb RN, Aung T, Medeiros FA. The Pathophysiology and Treatment of Glaucoma. JAMA. 2014;311(18):1901–1911. doi:10.1001/jama.2014.3192

5. Hayreh SS. Optic disc changes in glaucoma. Br J Ophthalmol. 1972;56(3):175–185. doi:10.1136/bjo.56.3.175

6. Hitchings RA, Spaeth GL. The optic disc in glaucoma. I: Classification. Br J Ophthalmol. 1976;60(11):778–785. doi:10.1136/bjo.60.11.778

7. Garway-Heath DF, Ruben ST, Viswanathan A, Hitchings RA. Vertical cup/disc ratio in relation to optic disc size: its value in the assessment of the glaucoma suspect. Br J Ophthalmol. 1998;82(10):1118–1124. doi:10.1136/bjo.82.10.1118

8. Foster PJ. The definition and classification of glaucoma in prevalence surveys. Br J Ophthalmol. 2002;86(2):238–242. doi:10.1136/bjo.86.2.238

9. Gordon MO, Beiser JA, Brandt JD, et al. The ocular hypertension treatment study: Baseline factors that predict the onset of primary open-angle glaucoma. Arch Ophthalmol. 2002;120(6):714–720.

10. Tielsch JM, Katz J, Quigley HA, Miller NR, Sommer A. Intraobserver and Interobserver Agreement in Measurement of Optic Disc Characteristics. Ophthalmology. 1988;95(3):350–356. doi: 10.1016/S0161-6420(88)33177-5

11. Varma R, Steinmann WC, Scott IU. Expert Agreement in Evaluating the Optic Disc for Glaucoma. Ophthalmology. 1992;99(2):215–221. doi:10.1016/S0161-6420(92)31990-6

12. LeCun Y, Bengio Y, Hinton G. Deep learning. Nature. 2015;521(7553):436–444. doi:10.1038/nature14539

13. Schmidt-Erfurth U, Sadeghipour A, Gerendas BS, Waldstein SM, Bogunović H. Artificial intelligence in retina. Prog Retin Eye Res. 2018;67(July):1–29. doi:10.1016/j.preteyeres.2018.07.004

14. Gulshan V, Peng L, Coram M, et al. Development and Validation of a Deep Learning Algorithm for Detection of Diabetic Retinopathy in Retinal Fundus Photographs. JAMA. 2016;316(22):2402. doi:10.1001/jama.2016.17216

15. Ting DSW, Cheung CY-L, Lim G, et al. Development and Validation of a Deep Learning System for Diabetic Retinopathy and Related Eye Diseases Using Retinal Images From Multiethnic Populations With Diabetes. JAMA. 2017;318(22):2211. doi:10.1001/jama.2017.18152

16. Brown JM, Campbell JP, Beers A, et al. Automated Diagnosis of Plus Disease in Retinopathy of Prematurity Using Deep Convolutional Neural Networks. JAMA Ophthalmol. 2018;136(7):803. doi:10.1001/jamaophthalmol.2018.1934

17. Burlina PM, Joshi N, Pekala M, Pacheco KD, Freund DE, Bressler NM. Automated Grading of Age-Related Macular Degeneration From Color Fundus Images Using Deep Convolutional Neural Networks. JAMA Ophthalmol. 2017;135(11):1170. doi:10.1001/jamaophthalmol.2017.3782

18. Burlina PM, Joshi N, Pacheco KD, Liu TYA, Bressler NM. Assessment of Deep Generative Models for High-Resolution Synthetic Retinal Image Generation of Age-Related Macular Degeneration. JAMA Ophthalmol. 2019;137(3):258. doi:10.1001/jamaophthalmol.2018.6156

19. Liu H, Wong DWK, Fu H, Xu Y, Liu J. DeepAMD: Detect Early Age-Related Macular Degeneration by Applying Deep Learning in a Multiple Instance Learning Framework. In: Asian Conference on Computer Vision.; 2019:625–640. doi:10.1007/978-3-030-20873-8_40

20. Zhao H, Li H, Maurer-Stroh S, Cheng L. Synthesizing retinal and neuronal images with generative adversarial nets. Med Image Anal. 2018;49:14–26. doi:10.1016/j.media.2018.07.001

21. Keel S, Wu J, Lee PY, Scheetz J, He M. Visualizing Deep Learning Models for the Detection of Referable Diabetic Retinopathy and Glaucoma. JAMA Ophthalmol. 2019;137(3):288. doi:10.1001/jamaophthalmol.2018.6035

22. Fu H, Cheng J, Xu Y, et al. Disc-Aware Ensemble Network for Glaucoma Screening From Fundus Image. IEEE Transactions on Medical Imaging. 2018;37(11):2493–2501. doi:10.1109/TMI.2018.2837012

23. Liu H, Li L, Wormstone IM, et al. Development and Validation of a Deep Learning System to Detect Glaucomatous Optic Neuropathy Using Fundus Photographs. JAMA Ophthalmol. 2019;137(12):1353. doi:10.1001/jamaophthalmol.2019.3501

24. Kumar JRH, Seelamantula CS, Kamath YS, Jampala R. Rim-to-Disc Ratio Outperforms Cup-to-Disc Ratio for Glaucoma Prescreening. Sci Rep. 2019;9(1):7099. doi:10.1038/s41598-019-43385-2

25. Cheng J, Liu J, Xu Y, et al. Superpixel Classification Based Optic Disc and Optic Cup Segmentation for Glaucoma Screening. IEEE Transactions on Medical Imaging. 2013;32(6):1019–1032. doi:10.1109/TMI.2013.2247770

26. Fu H, Cheng J, Xu Y, Wong DWK, Liu J, Cao X. Joint Optic Disc and Cup Segmentation Based on Multi-Label Deep Network and Polar Transformation. IEEE Transactions on Medical Imaging. 2018;37(7):1597–1605. doi:10.1109/TMI.2018.2791488

27. Jiang Y, Duan L, Cheng J, et al. JointRCNN: A Region-based Convolutional Neural Network for Optic Disc and Cup Segmentation. IEEE Transactions on Biomedical Engineering. 2020;67(2):335–343. doi: 10.1109/TBME.2019.2913211

28. Gu Z, Cheng J, Fu H, et al. CE-Net: Context Encoder Network for 2D Medical Image Segmentation. IEEE Transactions on Medical Imaging. 2019;38(10):2281–2292. doi:10.1109/TMI.2019.2903562

29. Orlando JI, Fu H, Barbosa Breda J, et al. REFUGE Challenge: A unified framework for evaluating automated methods for glaucoma assessment from fundus photographs. Med Image Anal. 2020;59:101570. doi:10.1016/j.media.2019.101570

30. Garway-Heath DF, Crabb DP, Bunce C, et al. Latanoprost for open-angle glaucoma (UKGTS): a randomised, multicentre, placebo-controlled trial. Lancet. 2015;385(9975):1295–1304. doi:10.1016/S0140-6736(14)62111-5

31. Falk T, Mai D, Bensch R, et al. U-Net: deep learning for cell counting, detection, and morphometry. Nat Methods. 2019;16(1):67–70. doi:10.1038/s41592-018-0261-2

32. Weerahandi S. Generalized Confidence Intervals. In: Exact Statistical Methods for Data Analysis. New York, NY: Springer New York; 1995:143–168. doi:10.1007/978-1-4612-0825-9_6

33. Efron B, Tibshirani R. Bootstrap Methods for Standard Errors, Confidence Intervals, and Other Measures of Statistical Accuracy. Stat Sci. 1986;1(1):54–75. doi:10.1214/ss/1177013815

34. Asaoka R, Murata H, Hirasawa K, et al. Using Deep Learning and Transfer Learning to Accurately Diagnose Early-Onset Glaucoma From Macular Optical Coherence Tomography Images. Am JOphthalmol. 2019;198:136–145. doi:10.1016/j.ajo.2018.10.007

35. Thompson AC, Jammal AA, Berchuck SI, Mariottoni EB, Medeiros FA. Assessment of a Segmentation-Free Deep Learning Algorithm for Diagnosing Glaucoma From Optical Coherence Tomography Scans. JAMA Ophthalmol. 2020;27710:1–7. doi:10.1001/jamaophthalmol.2019.5983

36. Maetschke S, Antony B, Ishikawa H, Wollstein G, Schuman J, Garnavi R. A feature agnostic approach for glaucoma detection in OCT volumes. Grulkowski I, ed. PLoS One. 2019;14(7):e0219126. doi:10.1371/journal.pone.0219126

37. Fu H, Baskaran M, Xu Y, et al. A Deep Learning System for Automated Angle-Closure Detection in Anterior Segment Optical Coherence Tomography Images. Am J Ophthalmol. 2019;203:37–45. doi:10.1016/j.ajo.2019.02.028

38. Fu H, Xu Y, Lin S, et al. Angle-Closure Detection in Anterior Segment OCT Based on Multilevel Deep Network. IEEE Trains Cybern. 2019:1–9. doi:10.1109/TCYB.2019.2897162

39. Wang L, Nie D, Li G, et al. Benchmark on Automatic Six-Month-Old Infant Brain Segmentation Algorithms: The iSeg-2017 Challenge. IEEE Transactions on Medical Imaging. 2019;38(9):2219–2230. doi:10.1109/TMI.2019.2901712

40. Yu S, Xiao D, Frost S, Kanagasingam Y. Robust optic disc and cup segmentation with deep learning for glaucoma detection. Comput Med Imaging Graph. 2019;74:61–71. doi:10.1016/j.compmedimag.2019.02.005

41. Zheng Y, Stambolian D, O’Brien J, Gee J. Optic Disc and Cup Segmentation from Color Fundus Photograph Using Graph Cut with Priors. In: International Conference on Medical Image Computing and Computer Assisted Intervention.; 2013: 75–82.

